# Ischemic Stroke Burden from Dietary Risks in China: 1990-2019 GBD Study

**DOI:** 10.1101/2024.12.02.24318360

**Authors:** Kui Duan, Yongran Cheng, Mingwei Wang, Shang Peng, Min Zhang, Jifeng Wang, Lan Ye, Zucai Xu, Zhanhui Feng

## Abstract

**Background:** In China, economic growth and lifestyle shifts have made dietary risks a major public health issue for ischemic stroke. This study examines the link between six dietary risks and ischemic stroke in China from 1990 to 2019, using 2019 Global Burden of Disease (GBD) data.

**Methods:** Data from GBD 2019 was primarily derived from Chinese monitoring systems, and the KaiLuan study focused on trends in ischemic stroke mortality and disability-adjusted life years (DALYs) associated with six dietary risks. Age-period-cohort and join-point regression methods were utilized, accounting for age, period, and cohort effects.

**Results:** In 2019, deaths and DALYs attributable to six dietary risks were 283,083 and 7,082,849, respectively, marking a significant increase of 124.7% and 108.4% since 1990. Despite overall declines in mortality and DALY rates from 1990 to 2019 (annual average percentage change (AAPC): −0.5% for DALYs and −0.6% for deaths), ischemic stroke rates due to diets high in red meat (AAPC: 1.4% for DALYs and 1.3% for deaths). For five dietary risks, local drift curves indicated increasing trends with age, except for low fiber intake. DALYs rates peaked at 85–89 years for all except sodium, which peaked at 75–79 years. Mortality rates slightly increased under 79 years, but rose noticeably over 79. Men had higher numbers and rates of dietary risk-related strokes, but they experienced smaller declines than women.

**Conclusions:** The study reveals an increasing ischemic stroke burden due to high red meat diets and decreasing trends due to low vegetable intakes. Dietary risks associated with high sodium, low fruit, vegetable, and whole grain intakes led to increased numbers but decreased rates of ischemic stroke outcomes.

## Introduction

In 2019, stroke was the second leading cause of death globally, causing significant disability ^1^. It has become the leading cause of death and disability in China^2^. According to GBD 2019, the ASRs of IS increased by 34.7% in China^3^. Despite medical advances, the burden of IS remains severe in China^4^. In 2019, dietary risks were the second leading cause of deaths for women worldwide and the third for men^5^. The INTERSTROKE study revealed ten risk factors contributing to 90% of stroke occurrences^6^, with dietary risks being significant for IS prevention. Advocating a healthy diet, emphasizing low salt and red meat intake while promoting fruits, vegetables, fiber, and whole grains, may reduce IS risks and promote overall health.

China’s economic and lifestyle changes have led to increased sugar and saturated fat consumption^7^. Previous studies have addressed some aspects of IS burden and risk factors in China, but comprehensive quantitative analysis of time, age, and gender trends in IS mortality and DALYs due to dietary risks is lacking^3^.

The GBD 2019 study provides a comprehensive database for assessing global disease burden and risk factors^8^. Our study aims to analyze and compare the temporal trends in IS deaths and DALYs associated with dietary risks in China from 1990 to 2019 using GBD 2019 data, join-point regression, and age-period-cohort methods. We will examine differences in IS deaths, DALY numbers, and rates between genders. This study aims to provide insights into the changing trends of IS death and DALYs related to dietary risks to inform future public health policies on stroke prevention and control in China.

## Methods

### Data Sources

The GBD 2019 study data(http://ghdx.healthdata.org/gbdresults-tool), provided by the Institute for Health Indicators and Assessment, quantifies global health losses from diseases, injuries, and risk factors between 1990 and 2019. The study evaluates IS mortality and DALYs linked to six dietary risk factors: (a) identifying six dietary risk factors; (b) estimating relative risk via systematic reviews; (c) assessing age, gender, location, and year-specific dietary levels; (d) determining theoretical minimum risk exposure levels, and (e) calculating population attributable scores, deaths, and DALYs. IS is clinically defined as the rapid onset or death of brain dysfunction, confirmed via medical records, CT, and MRI^9^.

### Dietary risk factor definitions

In GBD 2019, dietary risk factors include excessive intake of red meat (≥ 18–27 g/day) and sodium (≥ 1–5 g/day), low intake of fiber (≤ 19–28 g/day),fruits (≤ 200–300 g/day), vegetables (≤ 290–430 g/day) and whole grains (≤ 100–150 g/day). These selections are based on the strength of associations with diseases, evidence quality, and data availability.

### Statistical Analysis

The average annual percentage change (AAPC), the annual percentage change for each segment, and the 95% CIs were estimated to indicate the direction and magnitude of the trends.A join-point regression model evaluated time trends in mortality and DALYs from 1990 to 2019, identifying significant segmented trends. Age group cohort analysis assesses the impact of age, period, and cohort effects on IS-related mortality and DALYs. This study focuses on key estimates, including local drift, longitudinal age curve, and period (cohort) RR. Data from 1990-2019 includes deaths and demographics of individuals aged 25-94.

Join-point analysis was conducted using Join-point v.4.8.0.1 (April 2020) from the National Cancer Institute (Rockville, MD). (https://analysistools.nci.nih.gov/apc/) was used for the age-period-cohort analysis.Data analysis and visualization were performed using R (4.0.2). The R packages employed in the present study included “factoextra”, “dplyr”, “tidyverse”, “ggplot”, and “stats.” p < 0.05 was considered to indicate statistical significance.

## Results

The numbers and rates of diet-related IS deaths and DALYs between 1990 and 2019 are in Table 1. In 2019, the total numbers of IS deaths and DALYs due to six dietary risks were 283,083 (95% CI [179,034-404,919]) and 7,082,849 (95% CI [4,722,649-9,757,131]), with males accounting for 62.2% and 60.3% of cases, respectively. The death and DALYs rates were 15.8 (95% CI [10.1-22.9]) per 100,000 population and 357.8 (95% CI [239.7-496.5]) per 100,000 population. Since 1990, the numbers of IS deaths reached 125,989 (95% CI [84,502-175,611]), while DALYs amounted to 3,399,002 (95% CI [2,374,241-4,418,754]), marking significant increases of 124.7% and 108.4%, respectively. Adjusting for population growth and aging, death and DALYs rates declined from 18.7 (95% CI [12.4-26.4]) to 15.8 (95% CI [10.1-22.9]) deaths per 100,000 population and from 412.8 (95% CI [284.5-557.8]) to 357.8 (95% CI [239.7-496.5]) DALYs per 100,000 population between 1990 and 2019.

**Table 1.** Number and rate of diet-related IS death and DALYs between 1990 and 2019.

| Outcome | Years | sex | Numbers | 95%CI |  | Rate | 95%CI |  |
| --- | --- | --- | --- | --- | --- | --- | --- | --- |
|  |  |  |  | Lower | Upper |  | Low | Upper |
| Deaths | 1990 | Both | 125,989 | 84,502 | 175,661 | 18.7 | 12.4 | 26.4 |
|  |  | Female | 54,217 | 34,788 | 78,403 | 15.1 | 9.5 | 21.8 |
|  |  | male | 71,772 | 46,623 | 101,917 | 23.7 | 15.2 | 34.1 |
|  | 2019 | Both | 283,083 | 179,034 | 404,919 | 15.8 | 10.1 | 22.9 |
|  |  | Female | 106,923 | 61,852 | 166,011 | 11.1 | 6.4 | 17.1 |
|  |  | male | 176,160 | 113,378 | 252,415 | 22.3 | 14.1 | 32.3 |
| DALYs | 1990 | Both | 3,399,002 | 2,374,241 | 4,518,754 | 412.8 | 284.5 | 557.8 |
|  |  | Female | 1,494,957 | 1,018,131 | 2,022,399 | 351.9 | 237.0 | 478.7 |
|  |  | male | 1,904,045 | 1,305,928 | 2,594,168 | 485.5 | 326.4 | 674.2 |
|  | 2019 | Both | 7,082,849 | 4,722,649 | 9,757,131 | 357.8 | 239.7 | 496.5 |
|  |  | Female | 2,815,230 | 1,747,429 | 4,044,503 | 274.1 | 170.5 | 394.5 |
|  |  | male | 4,267,619 | 2,855,931 | 5,854,865 | 455.3 | 304.6 | 627.2 |

CI indicates confidence interval;DALYs,disability-adjusted life years;IS, ischemic stroke.

The trends of changes in numbers and rates of IS deaths and DALYs due to six dietary risks are in Tables 2 and 3, Figures S1–S6. They show substantial growth in IS deaths and DALYs linked to a high red meat diet, rising from 19,070 in 1990 to 73,275 in 2019 (AAPC 1.3, 95% CI [1.1-1.5]) and from 572,124 in 1990 to 1,976,144 in 2019 (AAPC 1.4, 95% CI [1.2-1.6]). Deaths and DALYs associated with a high sodium diet increased from 70,206 in 1990 to 153,366 and from 1,892,458 in 1990 to 3,979,298 in 2019, while the rate trends decreased with an AAPC of −0.6 (95% CI [−0.8 to −0.4]) and −0.5 (95% CI [−0.6 to −0.3]), respectively.

**Table 2.** Trends of Rates of IS DALYs Attributable to six dietary risks From 1990 to 2019 in China Using Join-Point Regression.

|  | Both sexes |  |  |  | Men |  |  |  | Women |  |  |  |
| --- | --- | --- | --- | --- | --- | --- | --- | --- | --- | --- | --- | --- |
|  | Segment | Period | APC(95%CI) | P value | Segment | Period | APC(95%CI) | P value | Segment | Period | APC(95%CI) | P value |
| Diet high in red meat | 1 | 1990-1997 | 0.5(0.3 to 0.7) | <0.01 | 1 | 1990-1998 | 0.9(0.8 to 1.1) | <0.01 | 1 | 1990-1997 | 0.2(0.0 to 0.5) | 0.07 |
|  | 2 | 1997-2004 | 2.9(2.7 to 3.1) | <0.01 | 2 | 1998-2004 | 3.1(2.9 to 3.4) | <0.01 | 2 | 1997-2004 | 2.9(2.6 to 3.2) | <0.01 |
|  | 3 | 2004-2007 | -0.3(-1.4 to 0.8) | 0.55 | 3 | 2004-2007 | 0.1(-1.1 to 1.4) | 0.82 | 3 | 2004-2007 | -0.9(-2.6 to 0.8) | 0.25 |
|  | 4 | 2007-2011 | 2.8(2.3 to 3.3) | <0.01 | 4 | 2007-2011 | 4.0(3.4 to 4.6) | <0.01 | 4 | 2007-2010 | 1.5(-0.1 to 3.2) | 0.06 |
|  | 5 | 2011-2017 | 1.1(0.8 to 1.3) | <0.01 | 5 | 2011-2016 | 1.5(1.1 to 1.8) | <0.01 | 5 | 2010-2019 | 0.8(0.7 to 1.0) | <0.01 |
|  | 6 | 2017-2019 | 0.1(-0.8 to 1.1) | 0.82 | 6 | 2016-2019 | -0.3(-0.8 to 0.3) | 0.32 |  |  |  |  |
|  | AAPC | 1990-2019 | 1.4(1.2 to 1.6) | <0.01 | AAPC | 1990-2019 | 1.7(1.5 to 1.9) | <0.01 | AAPC | 1990-2019 | 1.1(0.8 to 1.3) | <0.01 |
| Diet high in sodium | 1 | 1990-1993 | -0.7(-1.2 to -0.2) | 0.01 | 1 | 1990-1998 | -0.9(-1.1 to -0.8) | <0.01 | 1 | 1990-1993 | -0.9(-1.6 to 0.2) | 0.01 |
|  | 2 | 1993-1998 | -1.3(-1.6 to -1.0) | <0.01 | 2 | 1998-2004 | 0.8(0.5 to 1.2) | <0.01 | 2 | 1993-1998 | -1.8(-2.3 to -1.4) | <0.01 |
|  | 3 | 1998-2004 | 1.0(0.8 to 1.3) | <0.01 | 3 | 2004-2007 | -2.6(-4.0 to -1.1) | <0.01 | 3 | 1998-2004 | 1.3(0.9 to 1.6) | <0.01 |
|  | 4 | 2004-2007 | -3.2(-4.2 to -2.1) | <0.01 | 4 | 2007-2012 | 1.4(0.9 to 1.9) | <0.01 | 4 | 2004-2007 | -3.8(-5.2 to -2.4) | <0.01 |
|  | 5 | 2007-2011 | 0.5(-0.1 to 1.0) | 0.07 | 5 | 2012-2019 | -0.6(-0.8 to -0.4) | <0.01 | 5 | 2007-2015 | -1.2(-1.4 to -1.0) | <0.01 |
|  | 6 | 2011-2019 | -0.5(-0.6 to -0.4) | <0.01 |  |  |  |  | 6 | 2015-2019 | 0.1(-0.4 to 0.6) | 0.76 |
|  | AAPC | 1990-2019 | -0.5(-0.6 to -0.3) | <0.01 | AAPC | 1990-2019 | -0.2(-0.4 to -0.1) | <0.01 | AAPC | 1990-2019 | -0.9(-1.1 to -0.7) | <0.01 |
| Diet low in fiber | 1 | 1990-1998 | -2.8(-2.9 to -2.6) | <0.01 | 1 | 1990-1998 | -2.8(-2.9 to -2.6) | <0.01 | 1 | 1990-1998 | -2.8(-3.0 to -2.6) | <0.01 |
|  | 2 | 1998-2004 | -0.6(-0.9 to -0.3) | <0.01 | 2 | 1998-2004 | -0.6(-0.9 to -0.2) | <0.01 | 2 | 1998-2001 | 0.0(-1.8 to 1.8) | 0.99 |
|  | 3 | 2004-2007 | -5.3(-6.7 to -3.8) | <0.01 | 3 | 2004-2007 | -4.6(-6.1 to -3.1) | <0.01 | 3 | 2001-2004 | -1.3(-3.2 to 0.5) | 0.14 |
|  | 4 | 2007-2011 | -3.2(-3.9 to -2.4) | <0.01 | 4 | 2007-2011 | -2.0(-2.8 to -1.2) | <0.01 | 4 | 2004-2007 | -5.4(-7.2 to -3.5) | <0.01 |
|  | 5 | 2011-2014 | -4.4(-5.9 to -2.9) | <0.01 | 5 | 2011-2019 | -3.8(-3.9 to -3.6) | <0.01 | 5 | 2007-2015 | -4.5(-4.7 to -4.2) | <0.01 |
|  | 6 | 2014-2019 | -3.4(-3.8 to -3.1) | <0.01 |  |  |  |  | 6 | 2015-2019 | -3.0(-3.7 to -2.4) | <0.01 |
|  | AAPC | 1990-2019 | -2.9(-3.2 to -2.7) | <0.01 | AAPC | 1990-2019 | -2.7(-2.9 to -2.5) | <0.01 | AAPC | 1990-2019 | -3.1(-3.5 to -2.8) | <0.01 |
| diet low in fruit | 1 | 1990-1998 | -1.1(-1.3 to -0.9) | <0.01 | 1 | 1990-1997 | -0.9(-1.1 to -0.7) | <0.01 | 1 | 1990-1998 | -1.4(-1.6 to -1.2) | <0.01 |
|  | 2 | 1998-2001 | 0.8(-0.9 to 2.4) | 0.34 | 2 | 1997-2004 | 0.0(-0.3 to 0.3) | 0.92 | 2 | 1998-2001 | 1.0(-0.8 to 2.8) | 0.24 |
|  | 3 | 2001-2004 | -0.6(-2.2 to 1.1) | 0.48 | 3 | 2004-2007 | -4.1(-5.7 to -2.4) | <0.01 | 3 | 2001-2004 | -0.7(-2.4 to 1.0) | 0.37 |
|  | 4 | 2004-2007 | -4.4(-6.0 to -2.8) | <0.01 | 4 | 2007-2012 | -0.4(-1.0 to 0.1) | 0.13 | 4 | 2004-2007 | -4.9(-6.6 to -3.2) | <0.01 |
|  | 5 | 2007-2019 | -1.9(-2.0 to -1.8) | <0.01 | 5 | 2012-2019 | -1.8(-2.0 to -1.5) | <0.01 | 5 | 2007-2015 | -3.1(-3.3 to -2.9) | <0.01 |
|  |  |  |  |  |  |  |  |  | 6 | 2015-2019 | -1.4(-2.0 to -0.8) | <0.01 |
|  | AAPC | 1990-2019 | -1.5(-1.8 to -1.3) | <0.01 | AAPC | 1990-2019 | -1.1(-1.3 to -0.9) | <0.01 | AAPC | 1990-2019 | -1.9(-2.2 to -1.6) | <0.01 |
| diet low in vegetables | 1 | 1990-1995 | -7.2(-7.9 to -6.6) | <0.01 | 1 | 1990-1995 | -6.8(-7.5 to -6.2) | <0.01 | 1 | 1990-1995 | -7.7(-8.3 to -7.2) | <0.01 |
|  | 2 | 1995-1998 | -11.5(-14.1 to 8.8) | <0.01 | 2 | 1995-1999 | -10.8(-12.1 to -9.5) | <0.01 | 2 | 1995-1999 | -11.7(-12.8 to -10.6) | <0.01 |
|  | 3 | 1998-2008 | -9.5(-9.7 to -9.3) | <0.01 | 3 | 1999-2008 | -8.8(-9.0 to -8.5) | <0.01 | 3 | 1999-2002 | -8.7(-10.9 to -6.3) | <0.01 |
|  | 4 | 2008-2015 | -4.8(-5.1 to -4.5) | <0.01 | 4 | 2008-2011 | -3.3(-5.4 to -1.2) | <0.01 | 4 | 2002-2008 | -10.3(-10.8 to -9.9) | <0.01 |
|  | 5 | 2015-2019 | -2.3(-2.8 to -1.9) | <0.01 | 5 | 2011-2014 | -4.8(-6.7 to -2.9) | <0.01 | 5 | 2008-2015 | -5.3(-5.5 to -5.0) | <0.01 |
|  |  |  |  |  | 6 | 2014-2019 | -3.5(-3.9 to -3.1) | <0.01 | 6 | 2015-2019 | -1.6(-2.0 to -1.2) | <0.01 |
|  | AAPC | 1990-2019 | -7.2(-7.5 to -6.9) | <0.01 | AAPC | 1990-2019 | -6.9(-7.2 to -6.5) | <0.01 | AAPC | 1990-2019 | -7.5(-7.8 to -7.2) | <0.01 |
| diet low in | 1 | 1990-1998 | -0.3(-.4 to -0.2) | <0.01 | 1 | 1990-1997 | -0.2(-0.4 to 0.0) | 0.10 | 1 | 1990-1998 | -0.6(-0.8 to -0.4) | <0.01 |
| whole | 2 | 1998-2001 | 1.9(1.0 to 2.9) | 0.06 | 2 | 1997-2004 | 1.3(1.0 to 1.6) | <0.01 | 2 | 1998-2001 | 2.2(0.4 to 4.0) | 0.02 |
| grains | 3 | 2001-2004 | 0.9(-0.02 to 1.9) | <0.01 | 3 | 2004-2007 | -2.4(-3.9 to -0.9) | <0.01 | 3 | 2001-2004 | 0.7(-1.1 to 2.5) | 0.41 |
|  | 4 | 2004-2007 | -2.9(-3.8 to -1.9 ) | <0.01 | 4 | 2007-2012 | 1.3(0.8 to 1.8) | <0.01 | 4 | 2004-2007 | -3.3(-5.0 to -1.5) | <0.01 |
|  | 5 | 2007-2011 | 0.1(-0.3 to 0.6) | 0.53 | 5 | 2012-2019 | -0.7(-0.9 to -0.5) | <0.01 | 5 | 2007-2015 | -1.2(-1.5 to -1.0) | <0.01 |
|  | 6 | 2011-2019 | -0.6(-0.7 to -0.5) | <0.01 |  |  |  |  | 6 | 2015-2019 | -0.1(-0.7 to 0.5) | 0.80 |
|  | AAPC | 1990-2019 | -0.2(-4.1 to 0.0) | <0.01 | AAPC | 1990-2019 | 0.1(-0.1 to 0.2) | 0.54 | AAPC | 1990-2019 | -0.6(-0.9 to -0.3) | <0.01 |

AAPC indicates average annual percentage change; APC, annual percentage change.

**Table 3.** Trends of Rates of IS Death Attributable to six dietary risks From 1990 to 2019 in China Using Join-Point Regression.

|  | Both sexes |  |  |  | Men |  |  |  | Women |  |  |  |
| --- | --- | --- | --- | --- | --- | --- | --- | --- | --- | --- | --- | --- |
|  | Segment | Period | APC(95%CI) | P value | Segment | Period | APC(95%CI) | P value | Segment | Period | APC(95%CI) | P value |
| diet high<br>in red meat | 1 | 1990-1997 | 0.9(0.6 to 1.1) | <0.01 | 1 | 1990-1997 | 1.1(0.9 to 1.3) | <0.01 | 1 | 1990-1997 | 0.5(0.2 to 0.8) | <0.01 |
|  | 2 | 1997-2004 | 3.2(3.0 to 3.5) | <0.01 | 2 | 1997-2004 | 3.1(2.8 to 3.3) | <0.01 | 2 | 1997-2004 | 3.4(3.1 to 3.8) | <0.01 |
|  | 3 | 2004-2007 | -1.0(-2.5 to 0.5) | 0.17 | 3 | 2004-2007 | 0.0(-1.4 to 1.4) | 0.99 | 3 | 2004-2007 | -2.0(-3.8 to 0.0) | 0.05 |
|  | 4 | 2007-2011 | 3.0(2.2 to 3.7) | <0.01 | 4 | 2007-2011 | 4.5(3.8 to 5.2) | <0.01 | 4 | 2007-2011 | 1.4(0.4 to 2.3) | <0.01 |
|  | 5 | 2011-2019 | 0.2(0.0 to 0.3) | 0.03 | 5 | 2011-2015 | 1.2(0.6 to 1.9) | <0.01 | 5 | 2011-2015 | -1.0(-1.9 to -0.1) | <0.01 |
|  |  |  |  | <0.01 | 6 | 2015-2019 | -0.7(-1.1 to -0.2) | <0.01 | 6 | 2015-2019 | 0.7(0.1 to 1.3) | <0.01 |
|  | AAPC | 1990-2019 | 1.3(1.1 to 1.5) | <0.01 | AAPC | 1990-2019 | 1.7(1.5 to 1.9) | <0.01 | AAPC | 1990-2019 | 0.9(0.6 to 1.2) | 0.02 |
| diet high<br>in sodium | 1 | 1990-1998 | -1.0(-1.2 to -0.9) | <0.01 | 1 | 1990-1998 | -0.8(-1.0 to -0.7) | <0.01 | 1 | 1990-1998 | -1.6(-1.8 to -1.4) | <0.01 |
|  | 2 | 1998-2004 | 1.3(1.0 to 1.6) | <0.01 | 2 | 1998-2004 | 1.0(0.6 to 1.4) | <0.01 | 2 | 1998-2004 | 1.6(1.2 to 2.1) | <0.01 |
|  | 3 | 2004-2007 | -3.5(-4.8 to -2.2) | <0.01 | 3 | 2004-2007 | -2.5(-4.1 to -0.9) | <0.01 | 3 | 2004-2007 | -4.7(-6.7 to -2.7) | <0.01 |
|  | 4 | 2007-2011 | 0.7(0.0 to 1.4) | 0.05 | 4 | 2007-2012 | 1.9(1.3 to 2.4) | <0.01 | 4 | 2007-2011 | -1.1(-2.1 to -0.1) | <0.01 |
|  | 5 | 2011-2019 | -1.1(-1.2 to -1.0) | <0.01 | 5 | 2012-2019 | -1.1(-1.4 to -0.9) | <0.01 | 5 | 2011-2015 | -2.7(-3.7 to -1.6) | <0.01 |
|  |  |  |  |  |  |  |  |  | 6 | 2015-2019 | -0.3(-0.9 to 0.4) | 0.41 |
|  | AAPC | 1990-2019 | -0.6(-0.8 to -0.4) | <0.01 | AAPC | 1990-2019 | -0.3(-0.5 to -0.1) | 0.01 | AAPC | 1990-2019 | -1.2(-1.5 to -0.9) | <0.01 |
| diet low<br>in fiber | 1 | 1990-1998 | -2.5(-2.7 to -2.4) | <0.01 |  | 1990-1998 | -2.5(-2.6 to -2.3) | <0.01 | 1 | 1990-1997 | -3.0(-3.2 to -2.7) | <0.01 |
|  | 2 | 1998-2004 | -0.2(-0.6 to 0.2) | 0.28 |  | 1998-2004 | -0.4(-0.8 to 0.0) | 0.03 | 2 | 1997-2004 | -0.2(-0.6 to 0.1) | 0.19 |
|  | 3 | 2004-2007 | -5.7(-7.3 to -4.0) | <0.01 |  | 2004-2007 | -4.4(-6.0 to -2.7) | <0.01 | 3 | 2004-2007 | -6.4(-8.5 to -4.2) | <0.01 |
|  | 4 | 2007-2011 | -2.8(-3.7 to -2.0) | <0.01 |  | 2007-2012 | -1.7(-2.3 to -1.2) | <0.01 | 4 | 2007-2011 | -4.1(-5.2 to -2.9) | <0.01 |
|  | 5 | 2011-2015 | -5.3(-6.1 to -4.4) | <0.01 |  | 2012-2019 | -4.7(-4.9 to -4.5) | <0.01 | 5 | 2011-2015 | -6.3(-7.4 to -5.2) | <0.01 |
|  | 6 | 2015-2019 | -4.0(-4.6 to -3.5) | <0.01 |  |  |  |  | 6 | 2015-2019 | -3.6(-4.4 to -2.8) | <0.01 |
|  | AAPC | 1990-2019 | -3.0(-3.3 to -2.8) | <0.01 | AAPC | 1990-2019 | -2.7(-2.9 to -2.5) | <0.01 | AAPC | 1990-2019 | -3.4(-3.7 to -3.1) | <0.01 |
| diet low<br>in fruits | 1 | 1990-1998 | -0.7(-0.9 to -0.5) | <0.01 |  | 1990-1997 | -0.4(-0.7 to -0.2) | <0.01 | 1 | 1990-1997 | -1.1(-1.5 to -0.8) | <0.01 |
|  | 2 | 1998-2001 | 1.5(-0.3 to 3.4) | 0.10 |  | 1997-2004 | 0.4(0.1 to 0.8) | 0.01 | 2 | 1997-2004 | 0.8(0.4 to 1.3) | <0.01 |
|  | 3 | 2001-2004 | -0.2(-2.0 to 1.7) | 0.84 |  | 2004-2007 | -3.7(-5.6 to -1.8) | <0.01 | 3 | 2004-2007 | -5.9(-8.4 to -3.3) | <0.01 |
|  | 4 | 2004-2007 | -4.7(-6.4 to -2.9) | <0.01 |  | 2007-2012 | 0.4(-0.2 to 1.0) | <0.01 | 4 | 2007-2011 | -2.5(-3.9 to -1.2) | <0.01 |
|  | 5 | 2007-2010 | -0.9(-2.7 to 0.9) | 0.30 |  | 2012-2019 | -2.5(-2.7 to -2.2) | <0.01 | 5 | 2011-2015 | -4.4(-5.6 to -3.1) | <0.01 |
|  | 6 | 2010-2019 | -2.5(-2.7 to -2.3) | <0.01 |  |  |  |  | 6 | 2015-2019 | -1.7(-2.6 to -0.8) | <0.01 |
|  | AAPC | 1990-2019 | -1.4(-1.8 to -1.1) | <0.01 | AAPC | 1990-2019 | -0.9(-1.2 to -0.7) | <0.01 | AAPC | 1990-2019 | -1.9(-2.2 to -1.5) | <0.01 |
| diet low<br>in vegetable<br>s | 1 | 1990-1995 | -6.0(-6.9 to -5.2) | <0.01 |  | 1990-1995 | -5.6(-6.4 to -4.9) | <0.01 | 1 | 1990-1994 | -6.1(-7.4 to -4.8) | <0.01 |
|  | 2 | 1995-1998 | -9.1(-12.6 to -5.6) | <0.01 |  | 1995-1999 | -8.2(-9.9 to -6.5) | <0.01 | 2 | 1994-1998 | -8.9(-10.8 to -6.9) | <0.01 |
|  | 3 | 1998-2004 | -7.0(-7.8 to -6.3) | <0.01 |  | 1999-2008 | -7.0(-7.3 to -6.6) | <0.01 | 3 | 1998-2004 | -6.9(-7.7 to -6.1) | <0.01 |
|  | 4 | 2004-2007 | -9.2(-12.2 to -6.1) | <0.01 |  | 2008-2011 | -2.9(-5.9 to 0.2) | <0.01 | 4 | 2004-2007 | -10.2(-13.4 to -6.9) | <0.01 |
|  | 5 | 2007-2015 | -5.5(-5.8 to -5.1) | <0.01 |  | 2011-2019 | -5.0(-5.2 to -4.7) | <0.01 | 5 | 2007-2015 | -6.4(-6.8 to -6.0) | <0.01 |
|  | 6 | 2015-2019 | -3.6(-4.3 to -2.8) | <0.01 |  |  |  |  | 6 | 2015-2019 | -2.9(-3.7 to -2.0) | <0.01 |
|  | AAPC | 1990-2019 | -6.4(-6.9 to -5.9) | <0.01 | AAPC | 1990-2019 | -5.9(-6.3 to -5.6) | <0.01 | AAPC | 1990-2019 | -6.7(-7.2 to -6.3) | <0.01 |
| diet low | 1 | 1990-1997 | -0.1(-0.3 to 0.2) | 0.67 | 1 | 1990-1997 | 0.2(0.0 to 0.5) | 0.08 | 1 | 1990-1997 | -0.5(-0.8 to -0.1) | <0.01 |
| in whole<br>grains. | 2 | 1997-2004 | 1.8(1.4 to 2.1) | <0.01 | 2 | 1997-2004 | 1.6(1.3 to 1.9) | <0.01 | 2 | 1997-2004 | 1.9(1.5 to 2.4) | <0.01 |
|  | 3 | 2004-2007 | -3.4(-5.3 to -1.5) | <0.01 | 3 | 2004-2007 | -2.2(-4.0 to -0.4) | 0.02 | 3 | 2004-2007 | -4.2(-6.6 to -1.9) | <0.01 |
|  | 4 | 2007-2010 | 1.0(-1.0 to 3.0) | 0.32 | 4 | 2007-2012 | 1.9(1.3 to 2.5) | <0.01 | 4 | 2007-2011 | -0.8(-2.0 to 0.4) | 0.15 |
|  | 5 | 2010-2019 | -1.2(-1.4 to -1.0) | <0.01 | 5 | 2012-2019 | -1.4(-1.7 to -1.2) | <0.01 | 5 | 2011-2015 | -2.8(-4.0 to -1.6) | <0.01 |
|  |  |  |  |  |  |  |  |  | 6 | 2015-2019 | -0.4(-1.2 to 0.4) | 0.25 |
|  | AAPC | 1990-2019 | -0.2(-0.5 to 0.0) | <0.01 | AAPC | 1990-2019 | 0.2(0.0 to 0.4) | 0.13 | AAPC | 1990-2019 | -0.7(-1.0 to -0.3) | <0.01 |

AAPC indicates average annual percentage change; APC, annual percentage.

Furthermore, deaths due to a low-fiber diet increased from 12,918 in 1990 to 14,064 in 2019, while the rate trend declined (AAPC −3.0, 95% CI [−3.3 to −2.8]). Due to a low-fiber diet, DALYs decreased from 390,300 in 1990 to 371,032 in 2019, showing a similar rate decline (AAPC −2.9, 95% CI [−3.2 to −2.7]). The numbers caused by low vegetable intake also declined (deaths: 8,249 to 2,940; DALYs: 222,306 to 56,483), showing the greatest decline in DALYs (AAPC −7.2, 95% CI [−7.5 to −6.9]) and deaths (AAPC −6.4, 95% CI [−6.9 to −5.9]). Trends indicated rising numbers for low fruit (deaths: 19,854 to 34,191; DALYs: 568,842 to 838,299) and whole grains (deaths: 17,198 to 42,626; DALYs: 486,365 to 1,061,084), with the weakest DALY rate decline for the low whole grain diet (AAPC −0.2, 95% CI [−4.1 to 0.0]). IS deaths and DALYs in males were higher than in females, except for a low vegetable diet.

Our research indicated significant decreases within specific time frames. From 2004 to 2007, the greatest declines in IS DALY rates were observed for diets high in red meat (APC −0.3, 95% CI [−1.4 to 0.8]) and sodium (APC −3.2, 95% CI [−4.2 to −2.1]), low in fiber (APC −5.3, 95% CI [−6.7 to −3.8]), low in fruits, and low in whole grains. Between 1995 and 1998, the DALY rates for low-vegetarian diets decreased. Similar declining trends were observed in IS death rates, with women showing greater declines than men.

Figures 1 and 2 illustrate longitudinal age curves of IS DALY and death rates related to six dietary risks in China. Rates increased with age, peaking at 85–89 years for most populations, except for high-sodium diets, which peak at 75–79 years. In men, peaks occurred at 90–94 years, and in women, at 75–79 years. IS death rates slightly increased in those under 79, with noticeable rises in those over 79. Male IS death rates peaked at 90–94 years, except for low-fiber diets. In all age groups, men had higher DALY and death rates than women.

**Figure 1.**
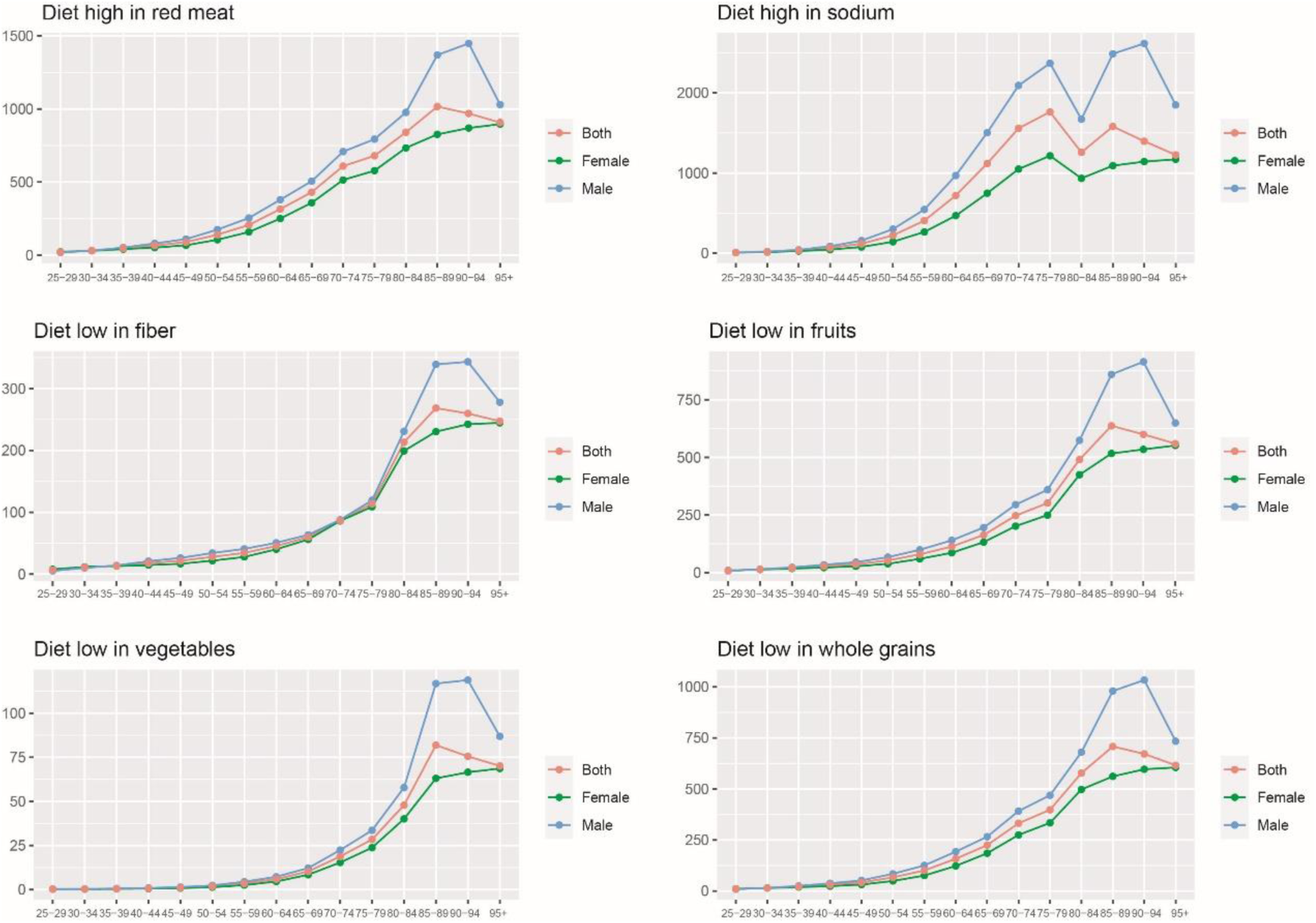
Trends in disability rates of IS caused by six different diets in different age groups in 2019.

**Figure 2.**
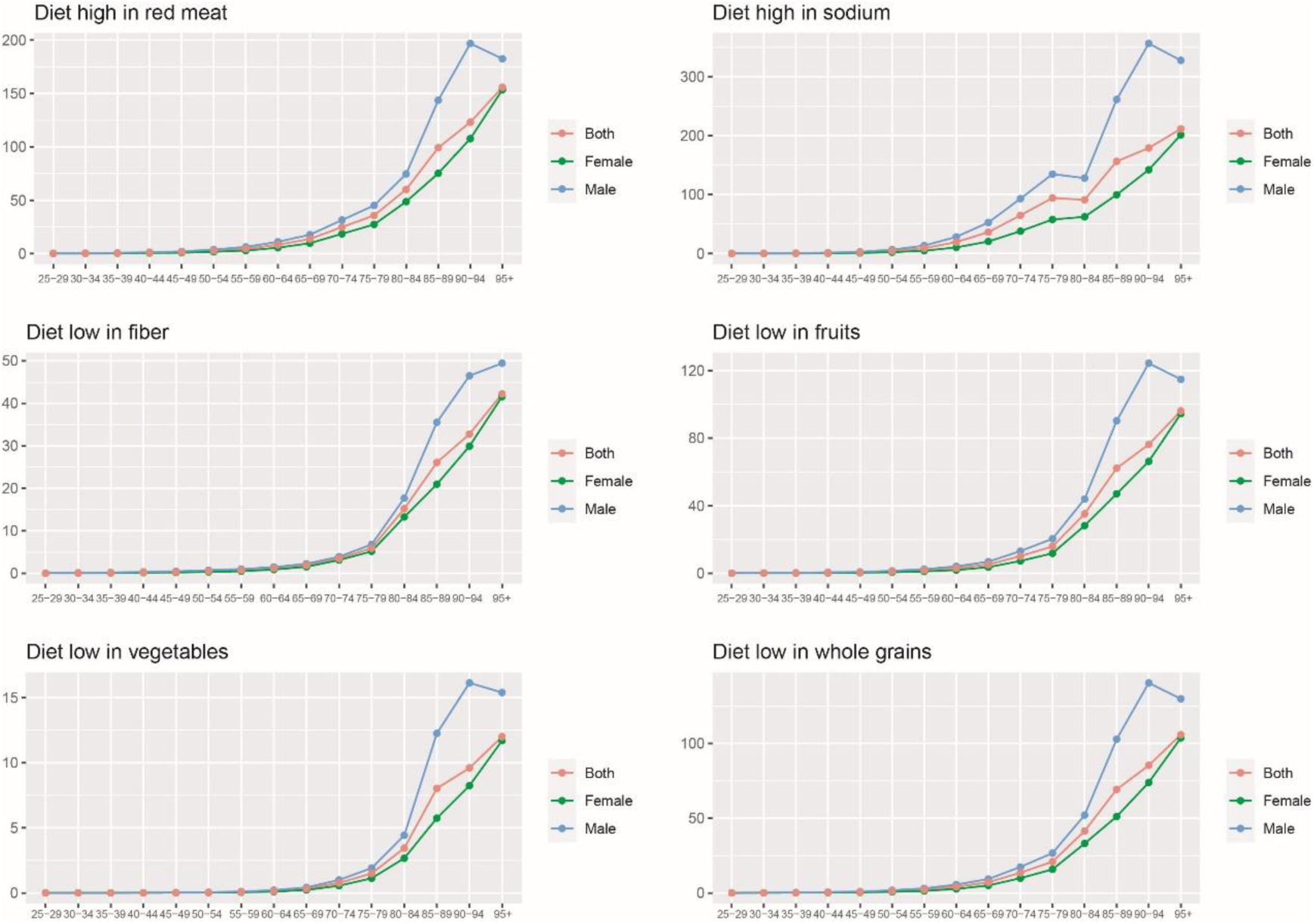
Trends in mortality rates of IS caused by six different diets in different age groups in 2019.

The changing trends of six dietary factors on IS risks in different age groups and genders from 1990 to 2019 are shown in Figures S7 to S12. We found IS risks from these dietary factors increased with age, being greater for men except for women under 35 before 2002. The impact of high-red meat consumption on IS in males increased from 1990 to 2019, while high-sodium diet risks weakened. From 2005 to 2019, the risks from high sodium and low fruit diets on IS increased for men aged 85 to 94. For men aged 80 to 94, the risks of low-grain diets on IS rose. The impacts of low-fiber, fruit, and vegetable diets on IS decreased, with almost no change in low-vegetarian diet impacts for people aged 44 to 79.

Age-period cohort analysis was used to calculate local drifts, estimating the AAPC of IS DALY and death rates in China attributed to six dietary risk factors across age groups, shown in Figure 3. The local drift curves for IS DALY rates of high red meat, low whole grains, high sodium, and low fruits displayed U-shaped patterns, with the most significant increases in those over 54. Low fiber’s IS DALY rates declined with age, peaking at 30-34, while low vegetable diet impacts increased with age. In the 30-44 age group, there was a significant decline in low-vegetarian diet-related IS DALYs, and in the 50-89 age group, there was a significant increase. The daily rate curves for men and women are similar to those in the general population. IS death rates associated with high red meat, sodium, low whole grain, and fruit diets showed W-shaped patterns, with the most significant decline in the 50–54 age group. Trends for both genders mirrored the overall population, with gradual increases among women with low fruit consumption and high red meat and sodium consumption.

**Figure 3.**
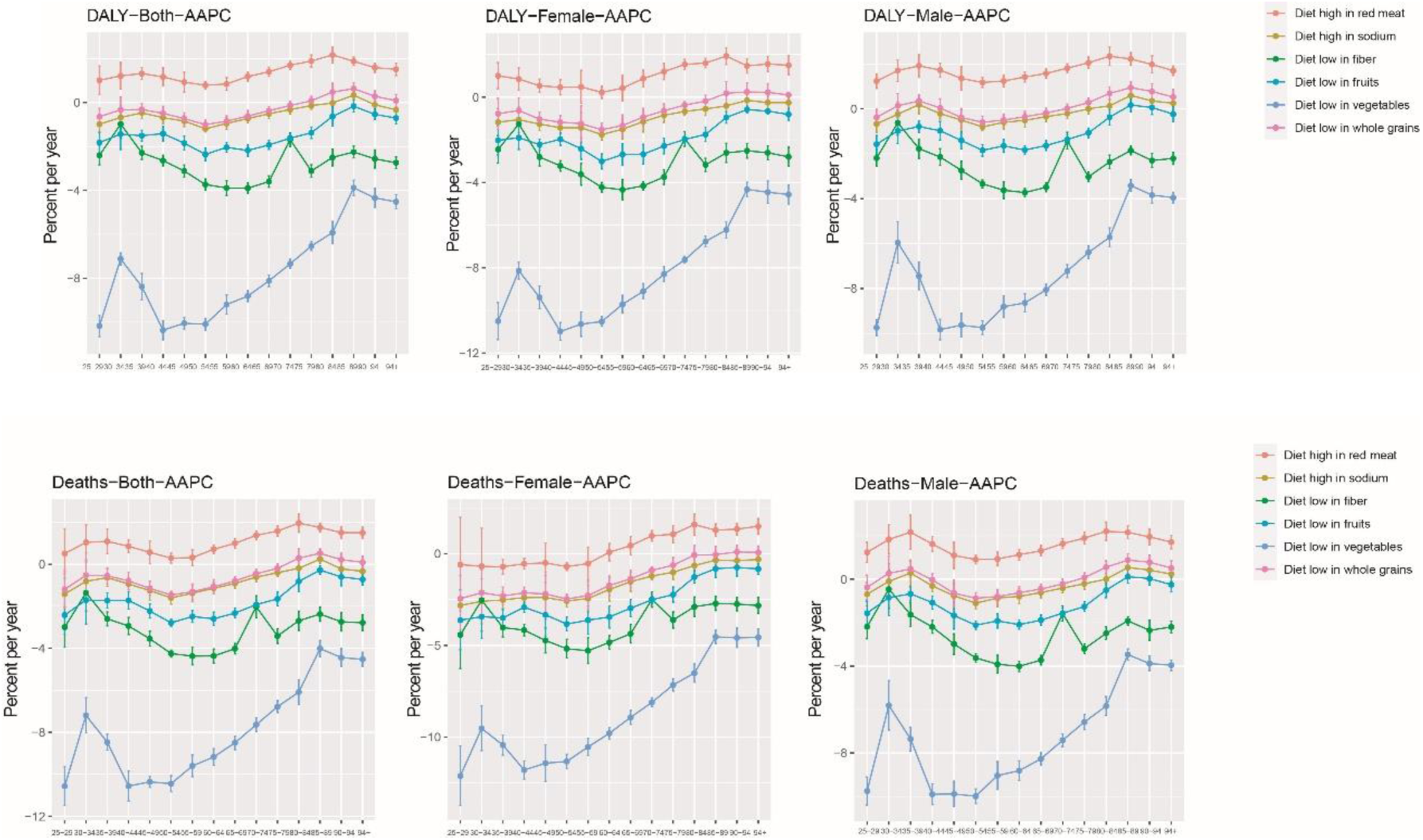
Trends in AAPC changes leading to mortality and disability in IS among 6 different diets at different age groups.AAPC indicates annual average percentage change;DALY,disability-adjusted life years.

## Discussion

This study, to the best of our knowledge, represents the first comprehensive analysis of cerebral IS attributable to six dietary risks in China from 1990 to 2019, based on the GBD study. In this study, we depicted the changes in deaths and DALYs in cerebral IS attributable to six dietary risks from 1990 to 2019 at the national level in China.

Using the GBD 2019 study, we found increases in the absolute numbers of IS deaths and DALYs in China due to high red meat and sodium diets and low consumption of fiber, fruits, and whole grains. Conversely, declining trends in IS deaths and DALYs related to low-fiber and vegetable diets were observed. Death and DALY rates attributed to the other five dietary risk factors also showed declines, particularly for low-fiber foods and vegetables. The increase in IS deaths and DALYs may be attributed to aging^10^, population growth, improved diagnostics, and unhealthy lifestyles. The decrease in rates over the past 30 years is likely due to better living conditions, improved medical care, and increased health awareness in China^11^.

It is widely acknowledged that age is a significant risk factor for IS^12^. Notably, IS deaths and DALYs due to six dietary factors increased dramatically with age, especially for high-sodium and low-fruit diets. However, we observed a decline in IS death rates among males over 94 years old, except for low-fiber diets. This is due to China’s accelerated aging and an increase in the elderly population. Men over 79 are more susceptible to other fatal diseases and tend to have unhealthy eating habits. Compared to younger patients, elderly IS patients have poorer outcomes due to more comorbidities, severe symptoms, medical complications, and fewer effective treatments^13^.

Gender significantly influences the IS disease burden^14^. In recent decades, IS deaths and DALYs in males were higher than in females, except for low vegetable diets. The rates of IS death and DALYs have consistently been higher in men. Females had lower negative AAPCs for IS death and DALY rates, indicating a greater threat to men. This may be due to the higher prevalence of unhealthy lifestyles among men ^15^ and the neuroprotective effect of estrogen in women ^16^. Men bear a heavier dietary risk burden and report less favorable attitudes and behavioral control regarding fruit and vegetable intake. Women generally have a lower hypertension incidence and adopt healthier lifestyles compared to men ^17^. If this trend continues, the gender gap is likely to widen.

A diet high in red meat has emerged as a significant risk factor for IS. Our study revealed a notable increase in IS-related deaths and DALYs associated with it, aligning with similar research findings ^18^. The most substantial decline in IS deaths and DALYs caused by it was within the 80–84 age group, suggesting older adults might have reduced their meat intake. However, an upward trend was noted among those aged 50–84 years, indicating interventions targeting this age group could yield significant health benefits. The numbers and DALYs related to it in men were higher than in women, as men prefer meat products. Prospective studies have shown a positive correlation between red meat consumption and IS risk ^19^, likely due to dietary saturated fatty acids, which increase IS risk ^20^.

We observed a notable increase in the number of IS deaths and DALYs attributed to a high-sodium diet, while the rates declined. An upward trend was observed in those aged 50 to 89 years, possibly due to higher sodium consumption among elderly adults. In 2009, Chinese adults over 50 consumed approximately 4.9 g/d (12.5 g/d) of sodium, while those under 50 consumed about 4.6 g/d (11.7 g/d) ^21^. Higher death and DALYs rates related to sodium were found in men compared to women, consistent with other studies ^22^.

A significant decrease in the number and rates of IS deaths and DALYs attributed to a low-vegetable diet was observed over the past 30 years, with a notable rise in the age group over 40 to 44 years. This suggests that individuals over the age of 40 should increase their vegetable consumption. Low intake of fruits and vegetables had been proven to be in relation to risk of ischemic stroke^23^, consistent with our findings. Fruits and vegetables provide fiber and micronutrients like potassium and folate, which help prevent strokes. They are also rich in dietary nitrates^24^, contributing to nitric oxide formation, lowering blood pressure, and offering vasoprotecting effects.

A slight decrease in the number of IS DALYs attributable to a low-fiber diet was noted, with an upward trend from 1998 to 2004. The death and DALYs rates associated with this diet significantly decreased from 1990 to 2019. In those over 65, an upward trend was observed, indicating the importance of increased fiber consumption in this age group. Fiber can reduce total lipids and LDL cholesterol levels^25^, and high fiber intake is associated with a reduced risk of diabetes^26^ and hypertension^27^, key IS risk factors.

Our study found a notable increase in the number of IS DALYs and deaths attributed to low whole grain intake over the past 30 years, especially in males, although the rates showed a slight decline. An upward trend was seen in those aged 50 to 84 years, indicating the need for increased whole grain consumption in adults over 50, particularly men, to prevent IS. Research supports our findings, suggesting that increased coarse grain consumption reduces IS risk and advocating for promoting whole grain consumption in China^28^. Whole grains provide richer nutrients than refined grains, reducing IS risk by regulating blood lipids and controlling blood pressure^29^. Encouraging whole grain intake aligns with the best dietary guidlines^30^.

In summary, the six dietary risk factors significantly impact IS incidence in China, posing a major public health concern. Our analysis revealed increases in IS deaths and DALYs due to high red meat diets, as well as decreases due to low vegetable diets. The other four dietary risks led to increased numbers but decreased rates of IS deaths and DALYs. The numbers and rates associated with these dietary risks have been consistently higher in men and the elderly compared to women and younger populations. In China, effective strategies to reduce sodium and red meat and increase fruit, vegetable, fiber, and whole grain intake should be implemented, particularly for stroke prevention in men and the elderly.

## Limitations

This study has several limitations. First, estimates were generated using GBD data and methodologies. Although generally reliable, GBD data has inherent limitations^31^. Second, dietary risks are defined and measured differently in the literature, making it difficult to accurately classify foods or nutrients. The GBD dataset’s dietary risks are not strictly classified, with overlaps in groups like whole grains, vegetables, fruits, and fiber. Third, the GBD framework does not classify ischemic stroke subtypes, preventing analysis of large artery atherosclerosis, cardioembolism, small vessel occlusion, and undetermined etiology^32^. Fourth, provincial-level analysis in China was not possible due to limited GBD 2019 data. Despite these limitations, this study provides valuable data for government measures to control dietary risk factors for ischemic stroke in China.

## Data Availability

The data that support the findings of this study are openly available in the GBD 2019 study data(http://ghdx.healthdata.org/gbdresults-tool).

## Nonstandard Abbreviations and Acronyms

AAPC: annual average percentage change
APC: annual percentage change
ASMRs: age-standardized mortality rates
ASRs: age-standardized rates
CI: confidence interval
CT: computed tomography
CVD: cardiovascular disease
DALYs: disability-adjusted life years
GBD: Global Burden of Disease
LDL: low density lipoprotein
MRI: magnetic resonance imaging
IS: ischemic stroke
RRs: rate ratios
WHO: World Health Organization

## Supplementary Material

Refer to Web version on PubMed Central for supplementary material.

## Acknowledgments

This study used publicly available deidentifying data from the 2019 Global Burden of Disease (GBD) provided by the Institute of Health Metrics and Assessment.

